# Mitochondrial respiratory activity and DNA damage in peripheral blood mononuclear cells in borderline personality disorder

**DOI:** 10.1101/2025.01.29.25321066

**Authors:** Alexander Behnke, Manuela Rappel, Laura Ramo-Fernández, R. Nehir Mavioğlu, Benjamin Weber, Felix Neuner, Ellen Bisle, Matthias Mack, Peter Radermacher, Stephanie H. Witt, Christian Schmahl, Alexander Karabatsiakis, Iris-Tatjana Kolassa

**Affiliations:** Clinical & Biological Psychology, Institute of Psychology and Education, Ulm University, D-89081 Ulm, Germany; Institute for Anesthesiologic Pathophysiology and Process Engineering, Ulm University Hospital, D-89081 Ulm, Germany; Department Genetic Epidemiology in Psychiatry, Central Institute of Mental Health, Medical Faculty Mannheim, Heidelberg University, D-68159 Mannheim, Germany; Department of Psychosomatic Medicine and Psychotherapy, Central Institute of Mental Health, Medical Faculty Mannheim, Heidelberg University, D-68159 Mannheim, Germany

## Abstract

Alterations in the central and peripheral energy metabolism are increasingly recognized as key pathophysiological processes in various psychiatric disorders. This case-control study investigates mitochondrial energy production and oxidative DNA damage in Borderline Personality Disorder (BPD). We compared mitochondrial respiration, density, and DNA damage in peripheral blood mononuclear cells between women with acute BPD, remitted BPD, and healthy controls (*n* = 32, 15, 29), matched for age and BMI. Acute BPD was characterized by reduced and less efficient mitochondrial ATP production compared to both remitted BPD and controls (e.g., coupling efficiency: *r*_x_ = −0.36 and −0.35, *p*_adj_’s < .037). Decreased mitochondrial activity was closely associated with greater DNA damage (e.g., coupling efficiency: *r*_S_ = −0.57, *p* < .001), although DNA damage did not differ between diagnostic groups. Our findings suggest mitochondrial energy production processes as promising and sensitive biomarkers for acute disorder severity and clinical remission in BPD.

## Introduction

Various psychiatric disorders are characterized by relevant changes in brain energy metabolism, with alterations in mitochondrial function and redox biology as major interfaces^1–6^. These brain alterations are complemented by peripheral metabolic abnormalities, including mitochondrial alterations in peripheral tissues that contribute to altered energy metabolism, oxidative stress, and chronic low-grade cytokine activity^6–13^. While mitochondrial alterations in both the brain and body are well-documented in conditions such as major depressive disorder (MDD), bipolar disorder (BP), and schizophrenia (SZ)^1,6^, research on the role of mitochondria in borderline personality disorder (BPD) remains limited.

BPD is a serious mental health condition characterized by emotional instability, inconsistent identity, interpersonal difficulties, feelings of emptiness, self-harming behavior, and an elevated risk of suicidal behavior^14^. The disorder shares symptoms with, and frequently co-occurs alongside, other psychiatric disorders such as MDD, BP, and posttraumatic stress disorder^14^, suggesting potential shared underlying pathophysiological mechanisms, including alterations in brain and body energy metabolism. However, the role of metabolic alterations in BPD remains largely unexplored^15^. Neuroimaging studies have provided initial evidence of reduced brain glucose metabolism and hemodynamic changes in BPD^16,17^, while studies of peripheral tissue such as blood and immune cells have identified reduced antioxidant cellular defense and elevated proinflammatory intracellular mediators, along with increased plasma cytokine activity in BPD^18–23^. To date, mitochondrial function in BPD has not been investigated.

Mitochondria are essential for various cellular processes^24^, including their major role in cellular energy metabolism through the production of adenosine triphosphate (ATP) via the tricarboxylic acid (TCA) cycle and oxidative phosphorylation (OxPhos). Alterations in mitochondrial dynamics and function, both in brain and periphery, have been observed in several psychiatric disorders, including MDD, BP, and SZ, and have been linked to several psychosocial risk factors for psychopathology^25–29^. In MDD, for example, studies in various peripheral tissues—such as skin and muscle cells, blood immune cells, and platelets—have established reduced basal OxPhos activity, decreased and less efficient ATP production, and alterations in mitochondrial content per cell. These alterations were consistently associated with the different symptom domains of MDD^7–10,30–35^. Based on these findings, we aimed to investigate whether mitochondrial alterations would contribute to symptom formation in BPD.

Moreover, mitochondria are a major source of reactive oxygen species (ROS) arising as by-products of OxPhos^36–38^. Oxidative stress occurs when ROS exceed a cell’s antioxidant capacity, causing damage to key cellular components, such as proteins, membranes, as well as nuclear and mitochondrial DNA^39,40^. There is a dynamic bidirectionality between oxidative damage and energy production in cells^39,41^. Cells tightly balance efficient mitochondrial ATP production against potentially harmful ROS induction using a dynamic regulation of protons leaking across the mitochondrial inner membrane^36–38^. Accumulating DNA damage activates DNA repair processes which may transiently decrease mitochondrial activity and biogenesis to prevent additional oxidative damage^39,40^.

High levels of DNA damage can lead to cell cycle dysregulation, chronic inflammation, and accelerated cellular aging^39,40^, mechanisms that are increasingly recognized as contributing to the pathophysiology of psychiatric disorders and their associated negative physical health outcomes^4,13,28^. There is consistent evidence of systemic oxidative stress in a range of psychiatric conditions, as indicated by elevated levels of oxidized DNA nucleotides in the cytoplasm, blood, and urine^28,42,43^, as well as more frequent DNA strand breaks in peripheral blood leukocytes^13,44–46^. However, in BPD, only preliminary evidence for increased oxidative DNA damage is currently available^22^.

In this case-control study, we investigate mitochondrial bioenergetics and DNA damage in BPD for the first time. We collected peripheral blood mononuclear cells (PBMCs) from three groups of age- and BMI-matched female participants: (i) 29 women with acute BPD, (ii) 15 women in remission from BPD, and (iii) 32 healthy controls. We hypothesized that individuals with acute BPD would exhibit reduced mitochondrial respiratory activity (e.g., less ATP production-related respiration) and elevated DNA damage compared to healthy controls. We also explored whether these alterations were associated with the current severity of the disorder or the lifetime diagnosis by comparing acute and remitted BPD. Lastly, we examined in the entire cohort whether mitochondrial bioenergetics were inversely related to oxidative DNA damage, as previously indicated^39^.

## Results

### Reduced mitochondrial energy production processes in BPD

Significant group differences were observed in mitochondrial ATP turnover (*F*(2,35.5) = 3.60, *p* = .038, η² = .167) and coupling efficiency (i.e., ATP turnover relative to basal respiration, χ²(2) = 6.89, *p* = .032, η²_rank_ = .067) (Table 2). *Post hoc* analyses revealed that individuals with acute BPD had lower ATP turnover (Cohen’s *d* = −0.62, *p*_adj_ = .037, Figure 1A) and less efficient ATP production (coupling efficiency: rank-biserial correlation *r*_x_ = −0.35, *p*_adj_ = .029, Figure 1C) compared to controls, indicating decreased mitochondrial energy production processes in acute BPD. Additionally, coupling efficiency was lower in acute BPD compared to remitted BPD (*r*_x_ = −0.36, *p*_adj_ = .037; Figure 1C). No significant group differences were found for leak respiration (Figure 1B), maximal respiration, spare respiratory capacity, and mitochondrial content in cells (Figure 1D) (Table 2).

**Figure 1.**
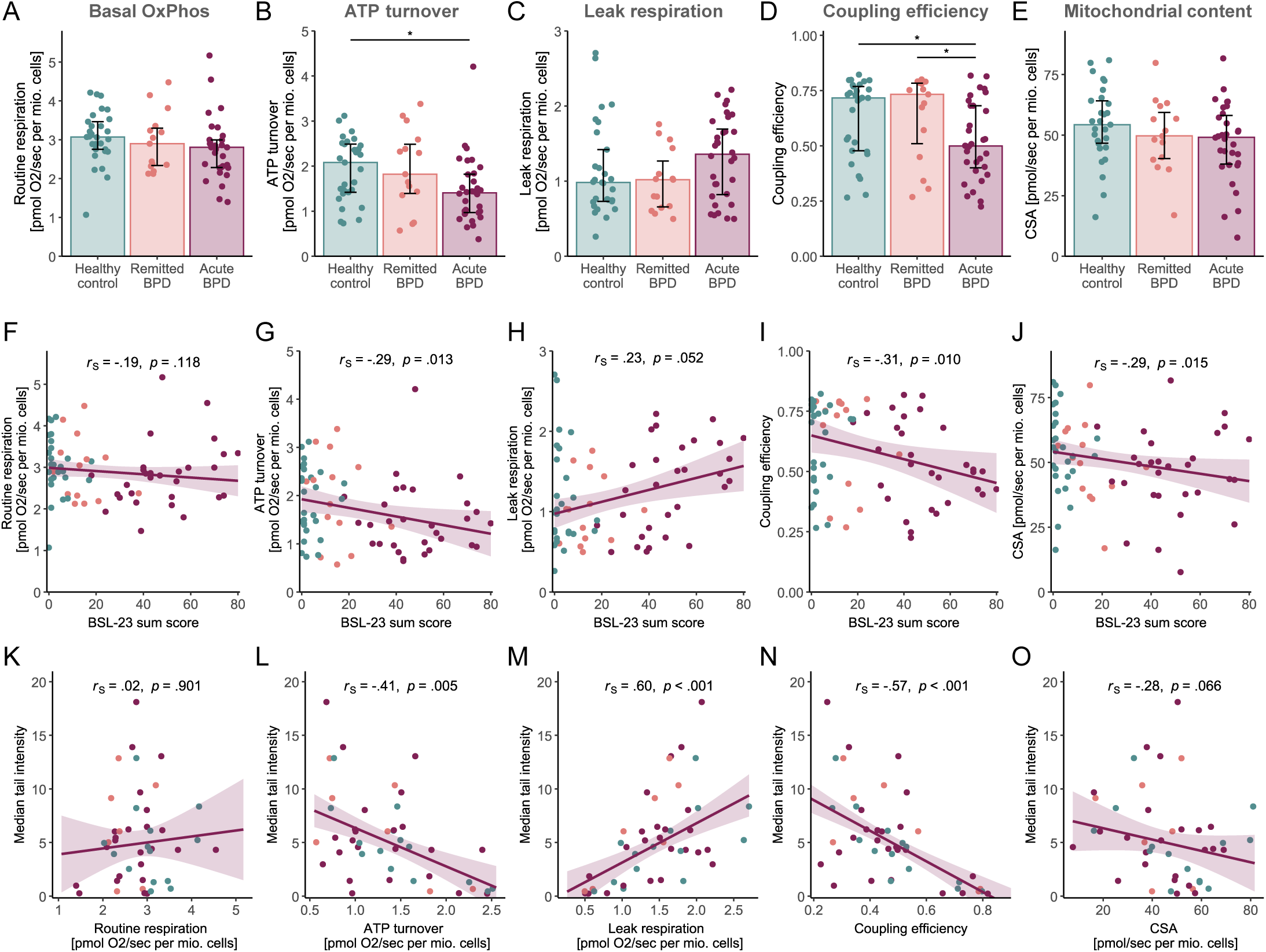
Group comparisons and correlations of mitochondrial parameters in peripheral blood mononuclear cells. (A-D) Bar plots represent median values of cellular respiration measures and cellular mitochondrial content, measured as citrate synthase activity (CSA), with interquartile ranges as error bars to depict differences between healthy controls (teal), remitted BPD (coral), and acute BPD (bordeaux). Data were analyzed with one-way Welch ANOVAs and Kruskal-Wallis tests as appropriate, with the significance of post hoc comparisons indicated (* *p*_adj_ < .050). (E-H) Scatter plots illustrate the association of self-reported BPD symptom severity with mitochondrial respiration measures and cellular mitochondrial content (CSA). (I-L) Scatter plots illustrate the associations of DNA damage (median tail intensity in the comet assay) with mitochondrial respiration measures and cellular mitochondrial content (CSA).

To rule out that the observed effects were influenced by comorbid episodes of MDD and/or antidepressant medication, we performed sensitivity analyses excluding such cases (Supplementary Table 2). These analyses confirmed the original findings and additionally revealed that individuals with acute BPD exhibited decreased basal OxPhos activity (routine respiration: *d* = −0.80, *p*_adj_ = .015) and lower mitochondrial content in cells (as indicated by citrate synthase activity: *r*_x_ = −0.43, *p*_adj_ = .017) compared to controls (Supplementary Figure 1). Notably, visual inspections of boxplots suggested that antidepressant medication might be confounding the original results, as acute BPD cases receiving antidepressant medication showed improved mitochondrial respiration compared to those without medication. Although this observation cannot be meaningfully statistically tested due to the low number of cases, the trend aligns with previous studies indicating that SSRIs may enhance mitochondrial respiration *in vitro*^47,48^.

### Symptom severity and mitochondrial function

Correlation analyses (Table 2) revealed that, regardless of diagnostic condition, higher BPD symptom severity was associated with reduced ATP turnover (*r*_S_ = −.29, *p* = .013, Figure 1E), lower coupling efficiency (*r*_S_ = −.31, *p* = .010, Figure 1G), and decreased mitochondrial content (*r*_S_ = −.29, *p* = .015, Figure 1H) in PBMCs. Greater symptom severity was marginally linked to higher leak respiration (*r*_S_ = .23, *p* = .052, Figure 1F). Sensitivity analyses (Supplementary Table 2) confirmed these findings by yielding numerically similar or even stronger associations, including a negative association between BPD symptom severity and basal OxPhos activity (i.e., routine respiration: *r*_S_ = −.39, *p* = .002).

We further explored the relationship between mitochondrial parameters and specific symptom domains in BPD, such as self-harming behavior, dissociative experiences, interpersonal difficulties, impulsivity, emotion dysregulation, trait anger, and depressed mood, as assessed by standardized clinical questionnaires. As summarized in Figure 2, most symptom domains showed a consistent pattern of negative correlations with markers of reduced mitochondrial energy metabolism (basal OxPhos activity, ATP turnover, coupling efficiency) and cellular mitochondrial content. The correlation pattern was more pronounced in the sensitivity subsample excluding cases with a comorbid MDD episodes and/or antidepressant medication.

**Figure 2.**
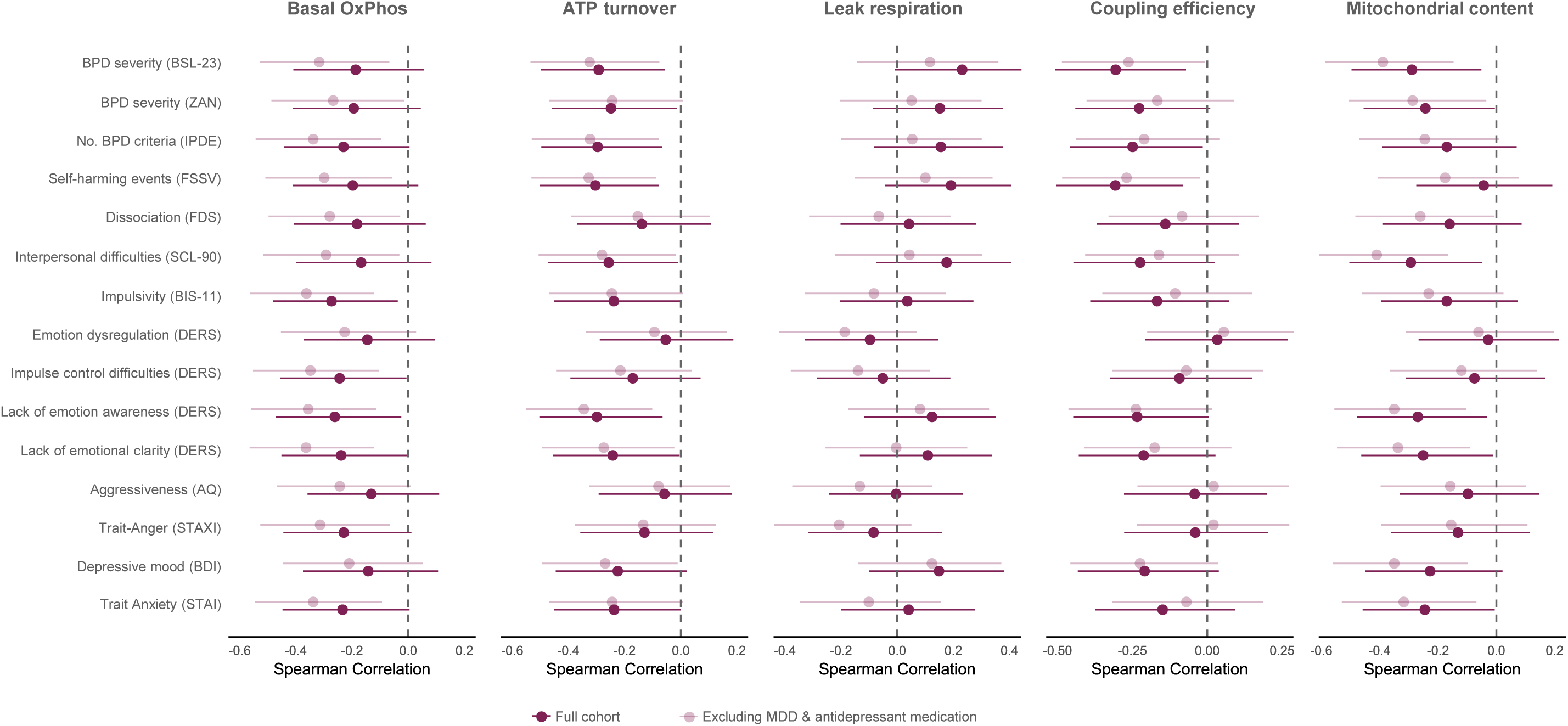
Forest plots illustrating explorative correlation analyses between mitochondrial parameters and measures of key symptom domains of Borderline Personality Disorder (BPD). Correlations were computed in the whole study cohort (bordeaux) and the sensitivity subsample excluding individuals with a comorbid major depression episode and/or receiving antidepressant medication (rosé). Dots and error bars represent effect sizes and 95% confidence intervals of Spearman correlations. Symptom domains were assessed with standardized clinical questionnaires detailed in the methods section.

### DNA damage and mitochondrial function

DNA damage levels did not significantly differ between groups (Kruskal-Wallis χ²(2) = 3.04, *p* = .219, η²_rank_ = .016; Table 2). However, there was a trend suggesting that higher BPD symptom severity was associated with greater DNA damage (*r*_S_ = .24, *p* = .054). We also tested whether DNA damage was inversely related to mitochondrial bioenergetics. In fact, DNA damage was negatively correlated with ATP turnover (*r*_S_ = −.41, *p* = .005, Figure 1I) and coupling efficiency (*r*_S_ = −.57, *p* < .001, Figure 1L), as well as mitochondrial content in trend (*r*_S_ = −.28, *p* = .066, Figure 1L). Additionally, DNA damage was positively correlated to mitochondrial leak respiration (*r*_S_ = .60, *p* < .001, Figure 1J).

Age and BMI were not associated to mitochondrial markers and DNA damage in the present study.

## Discussion

Individuals with acute BPD showed reduced mitochondrial energy production processes in PBMCs compared to controls, characterized by lower oxygen consumption devoted to ATP production. Furthermore, sensitivity analyses revealed significant reductions in basal OxPhos activity and cellular mitochondrial content in acute BPD when excluding concurrent MDD and/or antidepressant medication. Altogether, these findings—lower mitochondrial content, OxPhos activity, and ATP production—indicate a reduced energy supply of immune cells from mitochondria in BPD. Importantly, these alterations are not associated with the lifetime diagnosis of BPD but are instead linked to the current severity of the disorder. Accordingly, the bioenergetic changes were consistently correlated with key symptom domains of BPD, including self-harming behavior, interpersonal difficulties, impulsivity, emotion dysregulation, trait anger, and depressed mood. This pattern is consistent with findings in MDD and BP, further supporting the view that peripheral mitochondrial function alterations serve as a marker of acute disorder states and severity^1,6–13,45^.

We found no evidence of compromised mitochondrial maximal and reserve respiratory capacity in acute BPD. These findings suggest that the observed decrease in basal OxPhos activity and ATP production in BPD is not due to a ‘dysfunctional’ electron transport chain, but rather reflects a downregulation of mitochondrial activity and density. Such a regulatory decrease in OxPhos and mitochondrial density could serve multiple purposes, including the reduction of mitochondrial ROS induction. One mechanism for this regulation is an increase in proton leak across the inner mitochondrial membrane, which also lowers the efficiency of ATP production^36–38^. While our data did not show differences in mitochondrial leak between diagnostic groups, we found that increased leak respiration was associated with higher BPD symptom severity. This suggests that mitochondrial proton leak might be transiently upregulated with increasing symptom severity, rather than being a characteristic of the lifetime diagnosis of BPD.

Balancing mitochondrial ATP production and ROS induction is critical to cellular survival. Oxidative stress can substantially damage vital cell components, including mitochondrial and nuclear DNA via base oxidization and DNA strand breaks^13,39,41^. Previous studies have reported increased markers of cellular and systemic oxidative stress and a diminished antioxidative capacity in BPD^18,21–23^, which aligns with findings of oxidative damage markers, such as increased DNA strand breaks, in other psychiatric disorders^13,44–46^. However, we did not observe higher DNA damage in acute or lifetime BPD, although increased DNA damage was associated with more severe BPD symptoms.

We did find significant associations between DNA integrity and mitochondrial bioenergetics. Cells with less DNA damage exhibited better mitochondrial efficiency and ATP production, while cells with greater DNA damage showed increased mitochondrial proton leak. This finding is consistent with well-established regulatory mechanisms in which mitochondrial coupling and biogenesis are downregulated in response to DNA damage, likely to reduce ROS induction and prevent further oxidative damage while the DNA is being repaired^36–38^. DNA repair processes, particularly those facilitated by poly(ADPribose) polymerase 1 (PARP1), can directly diminish mitochondrial activity due to competition for nicotinamide adenine nucleotide (NAD^+^), as well as indirectly influence mitochondrial biogenesis processes through complex transcriptional effects^39,49,50^. In par with this, we found a marginal negative association between DNA damage and mitochondrial content (*p* = .066), with lower mitochondrial content observed in cells from individuals with more severe BPD symptoms. Other studies indicate that mitochondrial regulation might also be relevant for telomeric DNA maintenance^30^. A recent longitudinal study in chronically stressed individuals highlighted that the association between chronic stress and prematurely shortened telomeres was explained by a stress-related decrease in mitochondrial OxPhos capacity, which predicted a reduction in telomerase activity^51^. These findings, along with our own results, underscore the need for further research into the regulatory relationship between DNA integrity and mitochondrial dynamics in the context of chronic stress, mental health, and aging.

In summary, our findings in BPD contribute to the growing body of evidence highlighting complex alterations in the energy metabolism of the body and brain across psychiatric disorders^1,6^. These changes in energy metabolism may result from chronic stress exposure^27,51–53^ and could serve as a central hub for the multifaceted biological alterations observed in psychiatric disorders—such as oxidative stress, DNA and telomere homeostasis, and as chronic cytokine activity^30,43,44,46,54,55^—that may even limit individual treatment responses^56^. Future research needs to advance our mechanistic understanding of energy metabolism changes in symptom formation and remission of mental health problems. Studies are warranted to characterize energy metabolic alterations at multiple levels of the biological system, for example, including *in vitro* studies exposing cells to energetic challenges via stress hormones or DNA damage^44,50,52^, as well as *in vivo* studies comparing the biological impacts of physical and psychosocial stress paradigms in patient and control populations^57,58^.

### Limitations

Our cross-sectional case-control study investigated mitochondrial bioenergetics and DNA integrity in BPD using a female-only cohort, which may limit the generalizability of our findings on non-female individuals. However, a recent meta-analysis^59^ found no evidence for sex-binary differences in mitochondrial parameters. Effects of diurnal rhythmicity and food intake cannot be ruled out as the time of blood collection and fasting were not standardized. Differences in PBMC composition might also influence the results as PBMC subtypes are known to vary in mitochondrial content and activity^9,60^. In our study, PBMC samples containing more lymphocytes, and memory T cells in particular, exhibited lower ATP turnover, higher leak respiration, and higher DNA damage (Supplementary Table 3). However, there were no systematic differences in PBMC composition between diagnostic groups (Supplementary Table 4). Future studies should aim to differentiate the abundance and metabolic profiles of immune cell types to further explore these factors. Finally, the cross-sectional design of our study does not allow to draw conclusions about the causal or temporal relationship of symptoms and biological alterations.

### Conclusions

In conclusion, our study provides the first evidence of reduced mitochondrial energy production in peripheral immune cells from individuals with acute BPD. These alterations in cellular energy metabolism are likely transient, reflecting the current severity of the disorder, and may normalize with clinical remission. Our findings contribute to the growing body of research suggesting mitochondrial alterations as a promising and sensitive biomarker for acute psychiatric conditions and their severity. Further investigation into the underlying causes and mechanisms of metabolic changes in both the brain and periphery will help inform innovative disease models and treatment approaches for severe psychiatric disorders.

## Methods

### Study cohort

The study enrolled three groups, matched for age and BMI: The first group consisted of 32 women diagnosed with acute BPD, who met five or more diagnostic criteria according to DSM-5^61^. The second group comprised 15 women in remission from BPD, who had a disorder history as per DSM-5 criteria, but only met a maximum of three DSM-5 criteria, excluding self-harming behavior, within two years prior to study participation (see Table 1 for the number of BPD criteria fulfilled). The third group consisted of 29 women without a history of severe mental and somatic disorders listed in the exclusion criteria. Participants were part of the study cohort of the DFG Clinical Research Unit 256 on BPD^62^, which recruited individuals with different mental disorders and controls at the Central Institute of Mental Health (CIMH) Mannheim as well as the Dept. of Psychiatry, Heidelberg University, Germany.

**Table 1.** Sociodemographic and clinical characteristics of the study cohort.

|  | Controls (*n* = 29) | *Med (IQR)<sup>a</sup>* |  | Group comparisons |  |  |
| --- | --- | --- | --- | --- | --- | --- |
|  |  | Remitted BPD (*n* = 15) | Acute BPD (*n* = 32) | Test statistic<sup>b</sup> | *p* | Effect size |
| Age, years | 25.5 (9.8) | 32.0 (5.8) | 26.0 (12.0) | $\chi^2(2) = 3.52$ | .172 | .022 |
| Body mass index, kg/m<sup>2</sup> | 22.8 (3.7) | 21.5 (5.1) | 21.9 (6.6) | $\chi^2(2) = 0.42$ | .811 | .000 |
| Smoking, No. (%) | 0 (0) | 2 (13) | 0 (0) | $\chi^2(2) = 8.35$ | .038 | .332 |
| BPD symptoms (BSL-23) | 1.0 (4.5) | 12.5 (7.0) | 45.0 (19.2) | $\chi^2(2) = 55.78$ | <.001 | .791 |
| BPD criteria fulfilled (IPDE), No. | 0.0 (0.0) | 1.0 (2.8) | 6.0 (2.0) | $\chi^2(2) = 61.78$ | <.001 | .842 |
| Comorbid SCID diagnoses, No. | 0.0 (0.0) | (0.0) 0.5 | 1.5 (1.0) | $\chi^2(2) = 29.70$ | <.001 | .379 |
| Child maltreatment exposure (CTQ) | 29.0 (3.5) | 65.0 (15.0) | 57.0 (22.8) | $\chi^2(2) = 44.22$ | <.001 | .650 |
| Antidepressant medication, No. (%) | 0 (0) | 2<sup>c</sup> (13) | 7<sup>d</sup> (22) | $\chi^2(2) = 7.01$ | .016 | .304 |

<sup>a</sup> Descriptive statistic given in *Med (IQR)*, exception, count variables given in numbers and percentage.
<sup>b</sup> Data were analyzed with Kruskal-Wallis $\chi^2$ tests (effect size: $\eta^2_{\text{rank}}$ ) and Pearson $\chi^2$ tests for contingency tables (effect size: Cramer's *V*) as appropriate.
<sup>c</sup> Medication, No.: sertraline (1), escitalopram (1).
<sup>d</sup> Medication, No.: venlafaxine (1), citalopram and sertraline (1), fluoxetine (2), escitalopram (3).

**Table 2.** Group comparisons and correlations of mitochondrial parameters and DNA damage in peripheral blood mononuclear cells.

| Variable | Controls | Remitted BPD | Acute BPD | Group comparisons | | | Spearman correlation, $r_s$ ($p$) | |
| --- | --- | --- | --- | --- | --- | --- | --- | --- |
| | $M$ ($SD$), $n = 29$ | $M$ ($SD$), $n = 15$ | $M$ ($SD$), $n = 32$ | Test statistic<sup>a</sup> | $p$ | $\eta^2$ | BPD symptoms<sup>b</sup> | DNA damage<sup>c</sup> |
| Routine respiration<sup>d</sup> | 3.07 (0.68) | 2.94 (0.76) | 2.77 (0.79) | $F(2,37.4) = 1.23$ | .304 | .062 | -.19 (.118) | .02 (.901) |
| Leak respiration<sup>d</sup> | 1.14 (0.62) | 1.01 (0.41) | 1.31 (0.54) | $\chi^2(2) = 3.23$ | .199 | .017 | .23 (.052) | .60*** (<.001) |
| Maximal respiration<sup>d</sup> | 7.22 (2.50) | 7.08 (2.01) | 7.01 (2.67) | $F(2,41.4) = 0.05$ | .950 | .002 | -.02 (.875) | .20 (.190) |
| ATP turnover-related respiration<sup>d,e</sup> | 1.92 (0.70) | 1.93 (0.89) | 1.46 (0.73) | $F(2,35.5) = 3.60$ | .038* | .167 | -.29* (.013) | -.41** (.005) |
| Spare respiratory capacity<sup>d</sup> | 4.16 (2.04) | 4.14 (1.42) | 4.24 (2.11) | $F(2,43.4) = 0.02$ | .979 | .001 | .03 (.793) | .21 (.162) |
| Coupling efficiency<sup>f</sup> | 0.63 (0.18) | 0.63 (0.19) | 0.52 (0.17) | $\chi^2(2) = 6.89$ | .032* | .067 | -.31** (.010) | -.57*** (<.001) |
| Citrate synthase activity<sup>g</sup> | 54.30 (15.47) | 49.97 (15.05) | 46.94 (16.16) | $F(2,38.4) = 1.61$ | .213 | .077 | -.29* (.015) | -.28 (.066) |
| DNA damage<sup>c</sup> | 3.26<sup>i</sup> (3.33) | 5.35<sup>j</sup> (4.04) | 4.71 (4.38) | $\chi^2(2) = 3.04$ | .219 | .016 | .24 (.054) | – |

Notes. \* $p < .050$ , \*\* $p < .010$ , \*\*\* $p < .001$ , two-tailed.
<sup>a</sup> Data were analyzed with one-way Welch ANOVAs ( $F$ ) or Kruskal-Wallis tests ( $\chi^2$ , effect size: $\eta^2_{\text{rank}}$ ) as appropriate.
<sup>b</sup> Severity of BPD symptoms was evaluated using the self-report questionnaires 23-item Borderline Symptom List (BSL-23). See Figure 2 for additional symptom measures.
<sup>c</sup> Indicated as median tail intensity, i.e., the median of the percentage ratio of the fluorescence signal in the tail normalized to the fluorescence signal in the head of the comet assay indicating the % of the DNA in tail.
<sup>d</sup> in pmol O<sub>2</sub>/sec per million living cells.
<sup>e</sup> Games-Howell test for multiple comparisons of ATP turnover-related respiration between groups: acute BPD vs. controls, Cohen's $d = -0.62$ , $p_{\text{adj}} = .037$ ; acute vs. remitted BPD, $d = -0.62$ , $p_{\text{adj}} = .200$ ; remitted BPD vs. controls, $d = 0.01$ , $p_{\text{adj}} > .999$ .
<sup>f</sup> Conover tests for multiple comparisons of coupling efficiency between groups: acute BPD vs. controls, rank-biserial correlation $r_x = -0.35$ , $p_{\text{adj}} = .029$ ; acute vs. remitted BPD, $r_x = -0.36$ , $p_{\text{adj}} = .037$ ; remitted BPD vs. controls: $r_x = 0.04$ , $p_{\text{adj}} = .437$ .
<sup>g</sup> in pmol/s per million living cells. One missing value.
<sup>i</sup> Values of 3 cases missing due to insufficient cell material.
<sup>j</sup> Values of 4 cases missing due to insufficient cell material.

### Procedure

The CIMH recruited patients and controls through online advertisements. Study procedures followed the Declaration of Helsinki^63^ and were approved by the ethics committees of Heidelberg and Ulm University. Upon giving written informed consent, all participants underwent standardized clinical diagnostics performed by trained raters. BPD diagnoses were determined using the International Personality Disorder Examination interview (IPDE)^64^. Comorbid psychiatric disorders were assessed with the German Structured Clinical Interview for DSM-IV Axis-I Disorders (SCID-I)^65^ with criteria updated to DSM-5 (see Table 1 for details).

The following exclusion criteria applied for enrollment: current pregnancy; lifetime diagnoses of schizophrenia spectrum and other psychotic disorders; lifetime and current bipolar I disorder; drug and alcohol dependency during the past year; substance abuse during the past two months (excluding nicotine); epilepsy and major somatic diseases. All participants were required to be free of current use of psychotropic medication, except for individuals in the BPD groups who were allowed to continue their usual regime of antidepressant medication limited to SSRIs and SNRIs during study participation (Table 1). Furthermore, participants were not allowed to take on-demand medication such as tranquilizers and cortisone for up to three days prior to participation.

At the time of blood sampling, self-report questionnaires were used to assess the current severity of mental health symptoms. BPD symptoms from the past week were evaluated by the sum score of the 23-item Borderline Symptom List (BSL-23)^66^ and the Zanarini Rating Scale (ZAN-BPS)^67^. There was a high degree of concordance (*r*_S_ = .83–.89, *p*’s < .001) between the self-report (BSL-23, ZAN-BPS) and clinician-evaluated assessments (IPDE) of BPD.

Relevant risk factors and important symptom domains of BPD were assessed using standardized questionnaires, including the Childhood Trauma Questionnaire (CTQ)^68^ to record maltreatment, abuse, and neglect experienced between 0 to 18 years of age; the Beck’s Depression Inventory-II (BDI-II)^69^ to assess depressive symptoms; the State-Trait Anxiety Inventory (STAI)^70^ to measure trait anxiety; the Symptom Checklist-90-Revised (SCL-90-R)^71^ to broadly characterize psychopathological problems; the questionnaire to record self-harming behavior (FSVV)^72^; the State-Trait Anger Expression Inventory-2 (STAXI-2)^73^; the Buss and Perry Aggression Questionnaire (AQ)^74^ assessing the inclination to frustration, physical and verbal aggression, and interpersonal hostility/mistrust; the Barratt Impulsiveness Scale (BIS-11)^75^ measuring cognitive and motor impulsivity as well as lack of planning; the Difficulties in Emotion Regulation Scale (DERS)^76^ to record difficulties in recognizing and regulating negative emotions.

### Blood sampling and PBMC isolation

Approximately 30 ml of peripheral whole blood (non-fasting) was collected by venipuncture (Sarstedt, Nümbrecht, Germany) into EDTA-buffered collection tubes under sterile conditions between 10:00 a.m. and 3:00 p.m. during medical rounds. Shortly after collection, PBMCs were isolated from whole blood by Ficoll-Paque density gradient centrifugation according to the manufacturer’s protocol (GE Healthcare, Chalfont St. Giles, UK). Isolated PBMCs were washed three times with sterile phosphate-buffered saline (PBS; Invitrogen, USA) by centrifugation at 150g for 10 minutes at room temperature (Heraeus Megafuge, Thermo Fisher Scientific, USA). The resulting cell pellet was resuspended in ice-cold cryopreservation medium (dimethyl sulfoxide: Sigma-Aldrich, St. Louis, MO, USA; fetal calf serum (FCS): Sigma-Aldrich, USA; dilution 1:10; <5 million PBMC/ml) and stored at − 80°C for at least 6 hours in a prechilled isopropanol-filled cryocontainer (Nalgene, USA). Samples were transported in batches on dry ice to Ulm University.

### Mitochondrial activity

In accordance with good scientific and laboratory practice, all biological measurements described below were performed by one experimenter blinded to sample group assignment. PBMC aliquots were thawed, washed twice with pre-warmed PBS containing 2% FCS (Sigma Aldrich) and resuspended in 4 ml MiR-05 respiration medium. During pre-processing, 10 µl of the cell suspension was mixed with 10 µl of trypan blue staining solution to count the total number of cells and to estimate the percentage of dead cells in the sample for oxygen consumption rate correction (pmol O_2_/sec × million living cells) according to the manufacturer’s recommendation. Samples were measured in duplicate at 37°C using an O2k Oxygraph (Oroboros Instruments, Austria). Immediately after translocation into the O2k chambers, chambers were closed and 10 µl of sodium pyruvate (2 M stock, Sigma-Aldrich) was added. To profile respiratory states according to oxygen consumption, a standardized Substrate-Inhibitor-Uncoupler-Titration (SUIT) protocol was performed^10^. In short, it involves the sequential addition of oligomycin, FCCP, rotenone, and antimycin to record different mitochondrial functional states: routine respiration (R) was assessed as the oxygen consumption of unstimulated cells and serves as a measure of physiological basal OxPhos activity. By inhibiting complex V (ATP synthase), leak respiration (L) was measured as the residual respiration that compensates for proton leak, slippage, and cation cycling across the mitochondrial membrane^77^. Maximal respiration (M) represents the maximal capacity of the electron transport chain not limited by complex V and any proton gradients. Residual oxygen consumption of cells was determined after inhibition of all OxPhos-associated enzymes, and was subtracted from all raw values to correct for measures of cellular oxygen consumption not related to mitochondrial OxPhos and technical noise. L was subtracted from R to estimate ATP turnover-related respiration, and the difference between M and R represents the spare respiratory capacity of the system. Cellular oxygen flux was recorded in real time using DatLab software 6.1.0.7 (Oroboros Instruments). Respiration values were calculated as the mean of technical duplicates. Following the manufacturer’s description^77^, flux control ratios were calculated to normalize respiratory measurements for cell size and mitochondrial content, with the ATP turnover-related respiration relative to R indicating the coupling efficiency of ATP induction.

### Mitochondrial content of cells

The density of the intracellular mitochondrial network in PBMCs was assessed using the activity of the citrate synthase, a pacemaker enzyme in the TCA cycle^78^. After respirometry, one million living cells were collected, shock frozen with liquid nitrogen, and stored at −80°C until analysis. Citrate synthase activity (CSA) was measured spectrophotometrically in duplicates as previously described^10,53,79^.

### DNA damage

A cryopreserved PBMC aliquot was used for a single-cell gel electrophoresis assay (comet assay) under alkaline conditions^80,81^ to quantify DNA damage, including DNA single- and double-strand breaks as well as alkali-labile sites. Approximately one million cells per subject were thawed, washed in 9 ml PBS, and centrifuged at 1200 rpm for 10 min at 18°C (Heraeus Megafuge, Thermo Fisher Scientific). After supernatant removal, the cell pellet was resuspended with 10 µl PBS and separated into duplicate aliquots of approximately 75,000 cells each. Each aliquot was added to 120 µl of 0.5% low melting-point agarose (Sigma-Aldrich), and the resulting cell suspension was applied by submersion to microscope slides (Thermo Fisher Scientific) previously coated on one side with a layer of 1.5% medium electroendoosmosis agarose (Sigma-Aldrich).

Coated microscope slides were covered with a coverslip. After solidification (4 min at 4°C), the coverslip was removed and the slides were immersed in solution (2.5 M NaCl: Sigma-Aldrich, 100 mM EDTA, 10 mM Tris; p*H*: 10) with freshly added 1% Triton X-100 (Sigma-Aldrich) and 10% DMSO (Sigma-Aldrich) at 4°C for overnight lysis. The slides were then immersed in the electrophoresis tank containing alkaline buffer (300 mM NaOH: WVR Chemicals, 1mM Na2EDTA: AppliChem PanReac, Germany; p*H* > 13) for 40 min before electrophoresis at 25 V (0.7 V/cm; 300 mA) at 4°C for 40 min. After neutralization, the slides were washed three times with 0.4 M Tris-base (p*H* 7.5). To ensure technical quality, each electrophoresis assay included HeLa cells irradiated with 8 Gray (using a Crosslinker at 2 J/cm² at 365 nm) as a positive control and untreated HeLa cells as a negative control (data not shown). The slides were rinsed with distilled water, dehydrated in 99.8% ethanol for 5 min, and stored at room temperature until microscopy.

DNA was stained with 50 µl of ethidium bromide (10 mg/ml, Carl Roth, Germany) prior to fluorescence microscopy at 40x magnification (microscope BX41, Olympus Lifescience, Waltham, MA, USA; camera Basler scA1300 −32fm, Soda Vision, Singapore), equipped with a mercury vapor bulb with a 590 nm barrier filter and a 515-560 nm excitation filter; Zeiss, Germany). For each subject, 200 cells (100 from each duplicate slide) were randomly selected and analyzed using Comet Assay IV software (Instem, UK). We used the median tail intensity (% of the DNA in tail) to quantify DNA damage, defined as the percentage ratio of fluorescence signal in the tail normalized to the fluorescence signal in the head of the comet assay. Tail intensity is preferred over derived measurements (e.g., tail moment) for better inter-batch and inter-laboratory reliability^82^.

### Flow cytometry

Analysis of PBMC composition was done after high-resolution respirometry due to limited cell material. The cell suspension was removed from the O2k chamber for subsequent analysis of PBMC composition, focusing on preselected subpopulations. First, we used propidium iodide staining (Miltenyi Biotec, Germany) to separate live from dead cells using fluorescence-activated cell sorting (FACS). Second, antibodies separated helper T cells (CD3^+^CD4^+^) and cytotoxic T cells (CD3^+^CD8^+^) from total PBMCs. Third, CD45RA^+^ antibodies served distinguishing naïve cells from memory cells. FACS was performed using appropriate antibodies according to the manufacturer’s protocol (Miltenyi Biotech) on a FACSAria III cell sorter system (BD Biosciences, Germany). Quality controls of the separated lymphocyte subsets were performed on randomly selected samples to test the homogeneity of the isolated subsets. Raw data were processed using BD FACSDIVA software (BD Biosciences). Absolute numbers of viable lymphocytes (CD3^+^), helper T cells (CD3^+^CD4^+^), and cytotoxic T cells (CD3^+^CD8^+^) were counted, and total percentages were determined two-parameter fluorescence scattergrams by fluorescence intensity levels. The reported percentages of cells are relative to the total number of viable CD3^+^ cells (Supplementary Table 4).

### Statistical analysis

Statistical analyses were performed using R 4.1.3^83^. Bivariate associations were examined using Spearman correlations (*r*_S_). Groups were compared using one-way Welch ANOVAs and Kruskal-Wallis tests, as appropriate, followed by *post hoc* tests (Games-Howell, Conover). Family-wise error rates were adjusted using Tukey or Holm corrections. All analyses used α < .050, two-tailed, as the threshold of statistical significance.

## Data Availability

All data produced in the present study are available upon reasonable request to the authors

## Acknowledgements

We gratefully acknowledge the clinical staff of the Central Institute of Mental Health Mannheim for recruiting of patients and performing diagnostic interviews. We especially would like to thank Martin Jungkunz for the administrative support, Marija Gligorijević for the blood collection and logistics, as well as Slavica Radosavljević-Bjelić for the isolation and cryopreservation of PBMCs.

## Funding

The study was funded by institutional resources from I.-T. Kolassa. Clinical data and blood samples were obtained from the Clinical Research Unit 256 (Christian Schmahl, Grant No. SCHM 1526/13-1), which is funded by the German Research Foundation (Deutsche Forschungsgemeinschaft, DFG).

## Conflict of interest

The authors report no conflict of interest.

## Author contributions

ITK, MR, and AK designed the study. CS and SHW organized the data and specimen collection from patients within the DFG research unit 256. MR organized the study setup and data collection from healthy controls and performed clinical screenings as well as data entry with support of FN. AK and PR contributed expertise in PBMC processing, flow cytometry, and mitochondrial analyses. LRF, BW, and MR conduced the Comet assays under the supervision of AK and PR. Technical processing of biological measurements was supported by AB, EB, RNM, and FN. AB conducted the statistical analyses and interpreted the results together with MM, RNM, EB, and ITK. AB drafted the manuscript with critical input and revisions from all authors. ITK provided funding for this study and supervised all stages of the study.

## Supplementary Tables

**Supplementary Table 1.**
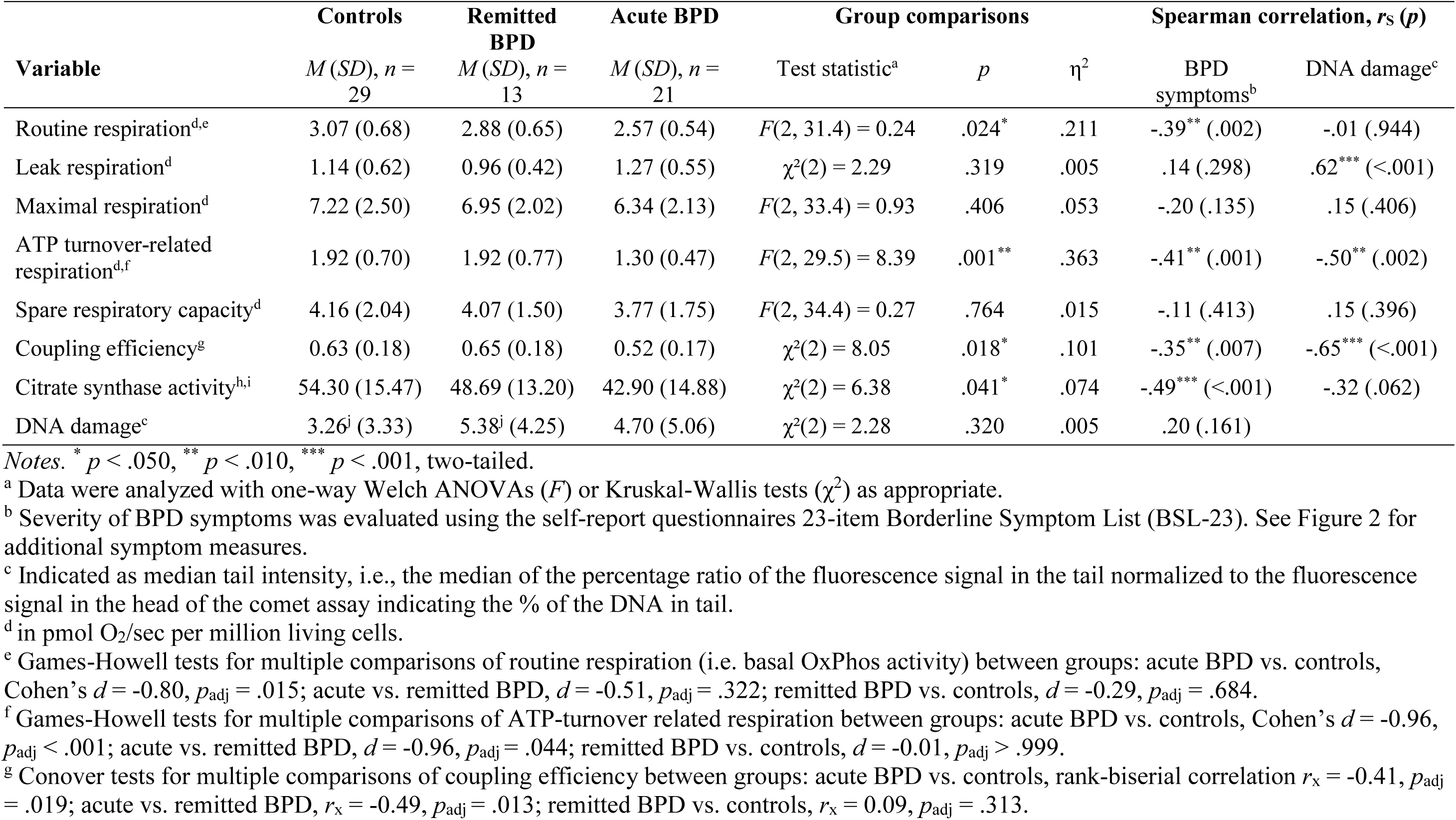
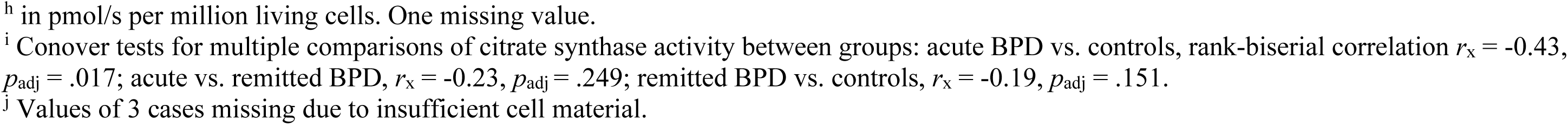
Sensitivity analysis of mitochondrial parameters and DNA damage in peripheral blood mononuclear cells under exclusion of cases with current major depression episode and/or antidepressant medication.

| Variable | Controls | Remitted BPD | Acute BPD | Group comparisons | | | Spearman correlation, $r_s$ ($p$) | |
| --- | --- | --- | --- | --- | --- | --- | --- | --- |
| | $M$ ($SD$), $n = 29$ | $M$ ($SD$), $n = 13$ | $M$ ($SD$), $n = 21$ | Test statistic<sup>a</sup> | $p$ | $\eta^2$ | BPD symptoms<sup>b</sup> | DNA damage<sup>c</sup> |
| Routine respiration<sup>d,e</sup> | 3.07 (0.68) | 2.88 (0.65) | 2.57 (0.54) | $F(2, 31.4) = 0.24$ | .024* | .211 | -.39** (.002) | -.01 (.944) |
| Leak respiration<sup>d</sup> | 1.14 (0.62) | 0.96 (0.42) | 1.27 (0.55) | $\chi^2(2) = 2.29$ | .319 | .005 | .14 (.298) | .62*** (<.001) |
| Maximal respiration<sup>d</sup> | 7.22 (2.50) | 6.95 (2.02) | 6.34 (2.13) | $F(2, 33.4) = 0.93$ | .406 | .053 | -.20 (.135) | .15 (.406) |
| ATP turnover-related respiration<sup>d,f</sup> | 1.92 (0.70) | 1.92 (0.77) | 1.30 (0.47) | $F(2, 29.5) = 8.39$ | .001** | .363 | -.41** (.001) | -.50** (.002) |
| Spare respiratory capacity<sup>d</sup> | 4.16 (2.04) | 4.07 (1.50) | 3.77 (1.75) | $F(2, 34.4) = 0.27$ | .764 | .015 | -.11 (.413) | .15 (.396) |
| Coupling efficiency<sup>g</sup> | 0.63 (0.18) | 0.65 (0.18) | 0.52 (0.17) | $\chi^2(2) = 8.05$ | .018* | .101 | -.35** (.007) | -.65*** (<.001) |
| Citrate synthase activity<sup>h,i</sup> | 54.30 (15.47) | 48.69 (13.20) | 42.90 (14.88) | $\chi^2(2) = 6.38$ | .041* | .074 | -.49*** (<.001) | -.32 (.062) |
| DNA damage<sup>c</sup> | 3.26<sup>j</sup> (3.33) | 5.38<sup>j</sup> (4.25) | 4.70 (5.06) | $\chi^2(2) = 2.28$ | .320 | .005 | .20 (.161) | |

Notes. \* $p < .050$ , \*\* $p < .010$ , \*\*\* $p < .001$ , two-tailed.
<sup>a</sup> Data were analyzed with one-way Welch ANOVAs ( $F$ ) or Kruskal-Wallis tests ( $\chi^2$ ) as appropriate.
<sup>b</sup> Severity of BPD symptoms was evaluated using the self-report questionnaires 23-item Borderline Symptom List (BSL-23). See Figure 2 for additional symptom measures.
<sup>c</sup> Indicated as median tail intensity, i.e., the median of the percentage ratio of the fluorescence signal in the tail normalized to the fluorescence signal in the head of the comet assay indicating the % of the DNA in tail.
<sup>d</sup> in pmol O<sub>2</sub>/sec per million living cells.
<sup>e</sup> Games-Howell tests for multiple comparisons of routine respiration (i.e. basal OxPhos activity) between groups: acute BPD vs. controls, Cohen's $d = -0.80$ , $p_{\text{adj}} = .015$ ; acute vs. remitted BPD, $d = -0.51$ , $p_{\text{adj}} = .322$ ; remitted BPD vs. controls, $d = -0.29$ , $p_{\text{adj}} = .684$ .
<sup>f</sup> Games-Howell tests for multiple comparisons of ATP-turnover related respiration between groups: acute BPD vs. controls, Cohen's $d = -0.96$ , $p_{\text{adj}} < .001$ ; acute vs. remitted BPD, $d = -0.96$ , $p_{\text{adj}} = .044$ ; remitted BPD vs. controls, $d = -0.01$ , $p_{\text{adj}} > .999$ .
<sup>g</sup> Conover tests for multiple comparisons of coupling efficiency between groups: acute BPD vs. controls, rank-biserial correlation $r_x = -0.41$ , $p_{\text{adj}} = .019$ ; acute vs. remitted BPD, $r_x = -0.49$ , $p_{\text{adj}} = .013$ ; remitted BPD vs. controls, $r_x = 0.09$ , $p_{\text{adj}} = .313$ .

**Supplementary Table 2.** Characterization of the cohort’s psychiatric comorbidity.

| | Controls (*n*=29) | No. (%) | | $\chi^2_{\text{Pearson}}(\text{df})$ | Test statistic | |
| --- | --- | --- | --- | --- | --- | --- |
|  |  | Remitted BPD (*n*=15) | Acute BPD (*n*=32) |  | *p*<sub>Fisher</sub> | Cramer's *V* |
| Major depressive disorder (F32.x/F33.x) | 0 (0) | 0 (0) | 6<sup>a</sup> (19) | 9.37 (2) | .014 | .358 |
| Dysthymic disorder (F34.1) | 0 (0) | 1 (7) | 7<sup>a</sup> (22) | 7.19 (2) | .015 | .333 |
| Adjustment disorder | 0 (0) | 0 (0) | 1 (3) | 1.39 (2) | .999 | .135 |
| Posttraumatic stress disorder | 0 (0) | 0 (0) | 10 (45) | 15.83 (2) | <.001 | .456 |
| Panic disorder | 0 (0) | 0 (0) | 8 (25) | 12.29 (2) | .002 | .402 |
| Agora phobia | 1 (3) | 1 (7) | 1 (3) | 0.37 (2) | .790 | .070 |
| Social phobia | 1 (3) | 0 (0) | 10 (31) | 12.66 (2) | .002 | .408 |
| Specific phobia | 1 (3) | 0 (0) | 2 (6) | 1.08 (2) | .999 | .119 |
| Obsessive-compulsive disorder | 3 (10) | 1 (7) | 1 (3) | 1.29 (2) | .508 | .130 |
| Bulimia nervosa | 0 (0) | 1 (7) | 5 (16) | 5.15 (2) | .077 | .260 |
| Binge eating disorder | 0 (0) | 0 (0) | 2 (6) | 2.82 (2) | .676 | .193 |
| Somatization disorder | 1 (3) | 0 (0) | 0 (0) | 1.64 (2) | .598 | .147 |
| Nicotine consumption | 0 (0) | 2 (13) | 0 (0) | 8.35 (2) | .038 | .332 |

*Note:* <sup>a</sup> including two cases with double depression (F33.x and F34.1). Diagnoses not listed in the table were not present among participants.

**Supplementary Table 3.** Bivariate associations of mitochondrial parameters and DNA damage assessed in bulk peripheral blood mononuclear cells (PBMCs) with the proportion of PBMC subpopulations.

|  | % CD3 T cells | % CD4 naïve T cells | % CD4 memory T cells | % CD8 naïve T cells | % CD8 memory T cells |
| --- | --- | --- | --- | --- | --- |
| Routine respiration | -.24* (.040) | -.12 (.306) | -.12 (.333) | -.25* (.037) | -.20 (.084) |
| Leak respiration | .39*** (<.001) | .26* (.028) | .64*** (<.001) | -.14 (.245) | .61*** (<.001) |
| Maximal respiration | -.10 (.405) | .12 (.302) | -.07 (.561) | -.01 (.943) | -.08 (.487) |
| ATP turnover-related respiration | -.50*** (<.001) | -.29* (.013) | -.55*** (<.001) | -.11 (.371) | -.59*** (<.001) |
| Spare respiratory capacity | -.02 (.883) | .21 (.073) | -.05 (.673) | .09 (.442) | -.03 (.776) |
| Coupling efficiency | -.50*** (<.001) | -.30* (.010) | -.65*** (<.001) | -.01 (.970) | -.65*** (<.001) |
| Citrate synthase activity | -.39*** (<.001) | -.29* (.014) | -.37** (.001) | -.21 (.078) | -.34** (.003) |
| DNA damage | .15 (.292) | .13 (.365) | .34* (.017) | -.15 (.314) | .31* (.026) |

Note: \* $p < .050$ , \*\* $p < .010$ , \*\*\* $p < .001$ , two-tailed significance of Spearman correlations ( $p$ -values in brackets). Cell counts represent percentage of cells types of total living PBMCs after respirometry.

**Supplementary Table 4.** Blood cell viability and composition of thawed immune cell samples.

|  | *M (SD)* |  |  | **Group comparisons** |  |  |
| --- | --- | --- | --- | --- | --- | --- |
| | Controls (*n* = 29) | Remitted BPD (*n* = 15) | Acute BPD (*n* = 32) | Welch ANOVA *F(df)* | *p* | $\eta^2$ |
| Viability of thawed cells in total PBMCs, % | 83.0 (7.4) | 85.7 (4.0) | 84.9 (6.3) | 1.14 (2, 42.69) | .329 | .051 |
| CD3 T cells, %<sup>a</sup> | 48.0 (12.3) | 47.0 (14.6) | 53.8 (15.8) | 1.47 (2, 36.45) | .242 | .075 |
| CD4 naïve T cells, %<sup>b</sup> | 17.1 (10.6) | 20.8 (15.2) | 18.8 (10.8) | 0.39 (2, 34.00) | .677 | .023 |
| CD4 memory T cells, %<sup>b,c</sup> | 15.9 (9.3) | 15.9 (10.2) | 21.1 (7.3) | 3.35 (2, 34.49) | .047 | .163 |
| CD8 naïve T cells, %<sup>b</sup> | 16.0 (6.3) | 14.8 (8.7) | 12.8 (4.8) | 2.30 (2, 32.52) | .116 | .124 |
| CD8 memory T cells, %<sup>b</sup> | 5.1 (4.1) | 5.1 (4.4) | 7.6 (5.1) | 2.50 (2, 37.69) | .096 | .117 |

<sup>a</sup> Percentage of cell types of total living PBMCs after respirometry.
<sup>b</sup> Percentage of total viable T cells (CD3<sup>+</sup>) after respirometry.
<sup>c</sup> Games-Howell tests for multiple comparisons of the percentage of CD4<sup>+</sup> memory T cells of total viable T cells (CD3<sup>+</sup>) after respirometry between groups: acute BPD vs. controls, Cohen's $d = -0.59$ , $p_{\text{adj}} = .059$ ; acute vs. remitted BPD, $d = -0.60$ , $p_{\text{adj}} = .206$ ; remitted BPD vs. controls, $d < 0.01$ , $p_{\text{adj}} > .999$ .

## Supplementary Figures

**Supplementary Figure 1.**
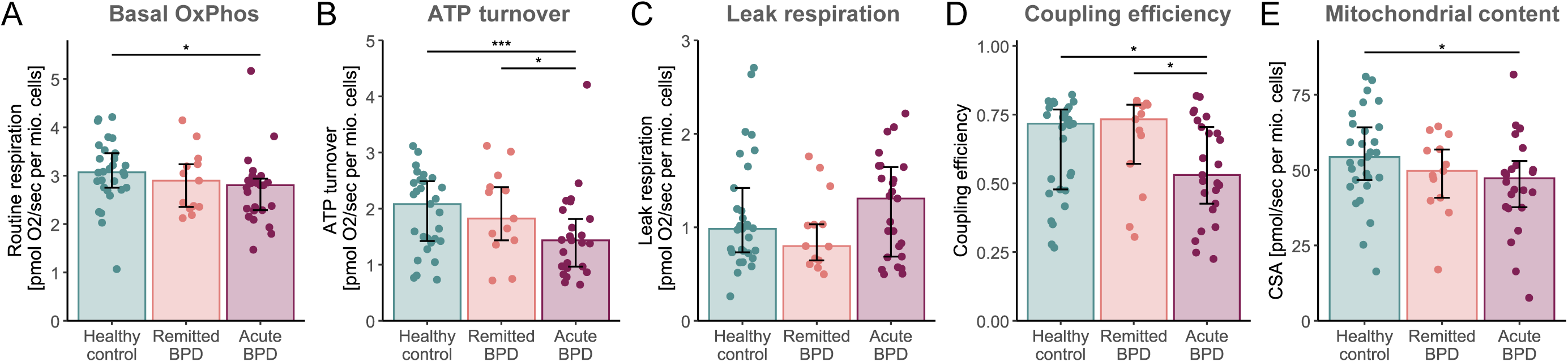
Comparisons of mitochondrial parameters in peripheral blood mononuclear cells between study groups under exclusion of cases with current major depression episode and/or antidepressant medication. (A-D) Bar plots represent median values of cellular respiration measures and cellular mitochondrial content, measured as citrate synthase activity (CSA), with interquartile ranges as error bars to depict differences between healthy controls (teal), remitted BPD (coral), and acute BPD (bordeaux). Data were analyzed with one-way Welch ANOVAs and Kruskal-Wallis tests as appropriate, with the significance of post hoc comparisons indicated (* *p*_adj_ < .050, *** *p*_adj_ < .001).

## Notes

### Competing Interest Statement

The authors have declared no competing interest.

### Author Declarations

Ethics committees of Ulm University and Heidelberg University gave ethical approval for this work.

## References

1 Ni P, Ma Y, Chung S. Mitochondrial dysfunction in psychiatric disorders. Schizophr Res 2024; 273: 62–77.

2 Henkel ND, Wu X, O’Donovan SM, Devine EA, Jiron JM, Rowland LM et al. Schizophrenia: a disorder of broken brain bioenergetics. Mol Psychiatry 2022; 27: 2393– 2404.

3 Mansur RB, Lee Y, McIntyre RS, Brietzke E. What is bipolar disorder? A disease model of dysregulated energy expenditure. Neurosci Biobehav Rev 2020; 113: 529–545.

4 Kim Y, Vadodaria KC, Lenkei Z, Kato T, Gage FH, Marchetto MC et al. Mitochondria, metabolism, and redox mechanisms in psychiatric disorders. Antioxid Redox Signal 2019; 31: 275–317.

5 Sarnyai Z, Ben-Shachar D. Schizophrenia, a disease of impaired dynamic metabolic flexibility: A new mechanistic framework. Psychiatry Res 2024; 342: 116220.

6 Papageorgiou MP, Filiou MD. Mitochondrial dynamics and psychiatric disorders: The missing link. Neurosci Biobehav Rev 2024; 165: 105837.

7 Gumpp AM, Behnke A, Bach AM, Piller S, Boeck C, Rojas R et al. Mitochondrial bioenergetics in leukocytes and oxidative stress in blood serum of mild to moderately depressed women. Mitochondrion 2021; 58: 14–23.

8 Triebelhorn J, Cardon I, Kuffner K, Bader S, Jahner T, Meindl K et al. Induced neural progenitor cells and iPS-neurons from major depressive disorder patients show altered bioenergetics and electrophysiological properties. Mol Psychiatry 2022. doi:10.1038/s41380-022-01660-1.

9 Gamradt S, Hasselmann H, Taenzer A, Brasanac J, Stiglbauer V, Sattler A et al. Reduced mitochondrial respiration in T cells of patients with major depressive disorder. iScience 2021; 24. doi:10.1016/j.isci.2021.103312.

10 Karabatsiakis A, Böck C, Kolassa S, Calzia E, Dietrich DE, Kolassa I-T. Mitochondrial respiration in peripheral blood mononuclear cells correlates with depressive subsymptoms and severity of major depression. Transl Psychiatry 2014; 4: e397.

11 Allen J, Romay-Tallon R, Brymer KJ, Caruncho HJ, Kalynchuk LE. Mitochondria and mood: Mitochondrial dysfunction as a key player in the manifestation of depression. Front Neurosci 2018; 12. doi:10.3389/fnins.2018.00386.

12 Giménez-Palomo A, Guitart-Mampel M, Meseguer A, Borràs R, García-García FJ, Tobías E et al. Reduced mitochondrial respiratory capacity in patients with acute episodes of bipolar disorder: Could bipolar disorder be a state-dependent mitochondrial disease? Acta Psychiatr Scand 2024; 149: 52–64.

13 Czarny P, Wigner P, Gałecki P, Śliwiński T. The interplay between inflammation, oxidative stress, DNA damage, DNA repair and mitochondrial dysfunction in depression. Prog Neuropsychopharmacol Biol Psychiatry 2018; 80: 309–321.

14 Bohus M, Stoffers-Winterling J, Sharp C, Krause-Utz A, Schmahl C, Lieb K. Borderline personality disorder. The Lancet 2021; 398: 1528–1540.

15 Saccaro LF, Schilliger Z, Dayer A, Perroud N, Piguet C. Inflammation, anxiety, and stress in bipolar disorder and borderline personality disorder: A narrative review. Neurosci Biobehav Rev 2021; 127: 184–192.

16 De La Fuente JM, Goldman S, Stanus E, Vizuete C, Morlán I, Bobes J et al. Brain glucose metabolism in borderline personality disorder. J Psychiatr Res 1997; 31: 531– 541.

17 Cattarinussi G, Delvecchio G, Moltrasio C, Ferro A, Sambataro F, Brambilla P. Effects of pharmacological treatments on neuroimaging findings in borderline personality disorder: A review of FDG-PET and fNIRS studies. J Affect Disord 2022; 308: 314–321.

18 Díaz-Marsá M, MacDowell KS, Guemes I, Rubio V, Carrasco JL, Leza JC. Activation of the cholinergic anti-inflammatory system in peripheral blood mononuclear cells from patients with Borderline Personality Disorder. J Psychiatr Res 2012; 46: 1610–1617.

19 Kahl KG, Rudolf S, Stoeckelhuber BM, Dibbelt L, Gehl H-B, Markhof K et al. Bone mineral density, markers of bone turnover, and cytokines in young women with borderline personality disorder with and without comorbid major depressive disorder. Am J Psychiatry 2005; 162: 168–174.

20 Kahl KG, Bens S, Ziegler K, Rudolf S, Dibbelt L, Kordon A et al. Cortisol, the cortisol-dehydroepiandrosterone ratio, and pro-inflammatory cytokines in patients with current major depressive disorder comorbid with borderline personality disorder. Biol Psychiatry 2006; 59: 667–71.

21 MacDowell KS, Díaz-Marsá M, Buenache E, Villatoro JML, Moreno B, Leza JC et al. Inflammatory and antioxidant pathway dysfunction in borderline personality disorder. Psychiatry Res 2020; 284: 112782.

22 Lee RJ, Gozal D, Coccaro EF, Fanning J. Narcissistic and borderline personality disorders: relationship with oxidative stress. J Personal Disord 2020; 34: 6–24.

23 Ruiz-Guerrero F, Gomez Del Barrio A, De La Torre-Luque A, Ayad-Ahmed W, Beato-Fernandez L, Polo Montes F et al. Oxidative stress and inflammatory pathways in female eating disorders and borderline personality disorders with emotional dysregulation as linking factors with impulsivity and trauma. Psychoneuroendocrinology 2023; 158: 106383.

24 Monzel AS, Enríquez JA, Picard M. Multifaceted mitochondria: moving mitochondrial science beyond function and dysfunction. Nat Metab 2023; 5: 546–562.

25 Trumpff C, Monzel AS, Sandi C, Menon V, Klein H-U, Fujita M, et al. Psychosocial experiences are associated with human brain mitochondrial biology. Proc Natl Acad Sci 2024; 121: e2317673121.

26 Picard M, Trumpff C, Burelle Y. Mitochondrial psychobiology: foundations and applications. Curr Opin Behav Sci 2019; 28: 142–151.

27 Gumpp AM, Boeck C, Behnke A, Bach AM, Ramo-Fernández L, Welz T et al. Childhood maltreatment is associated with changes in mitochondrial bioenergetics in maternal, but not in neonatal immune cells. Proc Natl Acad Sci 2020; 117: 24778–24784.

28 Giménez-Palomo A, Andreu H, de Juan O, Olivier L, Ochandiano I, Ilzarbe L et al. Mitochondrial dysfunction as a biomarker of illness state in bipolar disorder: A critical review. Brain Sci 2024; 14: 1199.

29 Gumpp AM, Behnke A, Ramo-Fernández L, Radermacher P, Gündel H, Ziegenhain U et al. Investigating mitochondrial bioenergetics in peripheral blood mononuclear cells of women with childhood maltreatment from post-parturition period to one-year follow-up. Psychol Med 2023; 53: 3793–3804.

30 Boeck C, Salinas-Manrique J, Calzia E, Radermacher P, von Arnim CAF, Dietrich DE et al. Targeting the association between telomere length and immuno-cellular bioenergetics in female patients with Major Depressive Disorder. Sci Rep 2018; 8. doi:10.1038/s41598-018-26867-7.

31 Gardner A, Boles RG. Beyond the serotonin hypothesis: Mitochondria, inflammation and neurodegeneration in major depression and affective spectrum disorders. Prog Neuropsychopharmacol Biol Psychiatry 2011; 35: 730–743.

32 Hroudová J, Fišar Z, Kitzlerová E, Zvěřová M, Raboch J. Mitochondrial respiration in blood platelets of depressive patients. Mitochondrion 2013; 13: 795–800.

33 Karabatsiakis A, Woike K, Behnke A, Kolassa I-T, C. Schönfeldt-Lecuona, Kiefer M et al. Testing the reversibility of impaired mitochondrial bioenergetic functioning in peripheral blood mononuclear cells from depressed patients by clinical-routine antidepressant treatment. In: Journal of Psychosomatic Research. 2020, p 110086.

34 Kuffner K, Triebelhorn J, Meindl K, Benner C, Manook A, Sudria-Lopez D et al. Major depressive disorder is associated with impaired mitochondrial function in skin fibroblasts. Cells 2020; 9: 884.

35 Zvěřová M, Hroudová J, Fišar Z, Hansíková H, Kališová L, Kitzlerová E et al. Disturbances of mitochondrial parameters to distinguish patients with depressive episode of bipolar disorder and major depressive disorder. Neuropsychiatr Dis Treat 2019; 15: 233–240.

36 Cheng J, Nanayakkara G, Shao Y, Cueto R, Wang L, Yang WY et al. Mitochondrial proton leak plays a critical role in pathogenesis of cardiovascular diseases. In: Santulli G (ed). Mitochondrial Dynamics in Cardiovascular Medicine. Springer International Publishing: Cham, 2017, pp 359–370.

37 Demine S, Renard P, Arnould T. Mitochondrial Uncoupling: A Key Controller of Biological Processes in Physiology and Diseases. Cells 2019; 8: 795.

38 Zhao R-Z, Jiang S, Zhang L, Yu Z-B. Mitochondrial electron transport chain, ROS generation and uncoupling (Review). Int J Mol Med 2019; 44: 3–15.

39 Fang EF, Scheibye-Knudsen M, Chua KF, Mattson MP, Croteau DL, Bohr VA. Nuclear DNA damage signalling to mitochondria in ageing. Nat Rev Mol Cell Biol 2016; 17: 308– 321.

40 Maynard S, Keijzers G, Gram M, Desler C, Bendix L, Budtz-Jørgensen E et al. Relationships between human vitality and mitochondrial respiratory parameters, reactive oxygen species production and dNTP levels in peripheral blood mononuclear cells. Aging 2013; 5: 850–864.

41 Kidane D, Chae WJ, Czochor J, Eckert KA, Glazer PM, Bothwell ALM et al. Interplay between DNA repair and inflammation, and the link to cancer. Crit Rev Biochem Mol Biol 2014; 49: 116–139.

42 Palta P, Samuel LJ, Miller ER, Szanton SL. Depression and oxidative stress: Results from a meta-analysis of observational studies. Psychosom Med 2014; 76: 12–19.

43 Jorgensen A, Baago IB, Rygner Z, Jorgensen MB, Andersen PK, Kessing LV et al. Association of Oxidative Stress–Induced Nucleic Acid Damage With Psychiatric Disorders in Adults: A Systematic Review and Meta-analysis. JAMA Psychiatry 2022; 79: 920–931.

44 Behnke A, Mack M, Fieres J, Christmann M, Bürkle A, Moreno-Villanueva M et al. Expression of DNA repair genes and its relevance for DNA repair in peripheral immune cells of patients with posttraumatic stress disorder. Sci Rep 2022; 12: 18641.

45 Czarny P, Kwiatkowski D, Kacperska D, Kawczyńska D, Talarowska M, Orzechowska A et al. Elevated level of DNA damage and impaired repair of oxidative DNA damage in patients with recurrent depressive disorder. Med Sci Monit 2015; 21: 412–418.

46 Morath J, Moreno-Villanueva M, Hamuni G, Kolassa S, Ruf-Leuschner M, Schauer M et al. Effects of psychotherapy on DNA strand break accumulation originating from traumatic stress. Psychother Psychosom 2014; 83: 289–297.

47 Cikánková T, Fišar Z, Hroudová J. In vitro effects of antidepressants and mood-stabilizing drugs on cell energy metabolism. Naunyn Schmiedebergs Arch Pharmacol 2020; 393: 797–811.

48 Fernström J, Mellon SH, McGill MA, Picard M, Reus VI, Hough CM et al. Blood-based mitochondrial respiratory chain function in major depression. Transl Psychiatry 2021; 11: 593.

49 Murata MM, Kong X, Moncada E, Chen Y, Imamura H, Wang P et al. NAD+ consumption by PARP1 in response to DNA damage triggers metabolic shift critical for damaged cell survival. Mol Biol Cell 2019; 30: 2584–2597.

50 Thomas M, Palombo P, Schuhmacher T, Von Scheven G, Bazylianska V, Salzwedel J et al. Impaired PARP activity in response to the β-adrenergic receptor agonist isoproterenol. Toxicol In Vitro 2018; 50: 29–39.

51 Guillen-Parra M, Lin J, Prather AA, Wolkowitz OM, Picard M, Epel ES. The relationship between mitochondrial health, telomerase activity and longitudinal telomere attrition, considering the role of chronic stress. Sci Rep 2024; 14: 31589.

52 Bobba-Alves N, Sturm G, Lin J, Ware SA, Karan KR, Monzel AS et al. Chronic Glucocorticoid Stress Reveals Increased Energy Expenditure and Accelerated Aging as Cellular Features of Allostatic Load. Cell Biology, 2022 doi:10.1101/2022.02.22.481548.

53 Boeck C, Koenig AM, Schury K, Geiger ML, Karabatsiakis A, Wilker S et al. Inflammation in adult women with a history of child maltreatment: The involvement of mitochondrial alterations and oxidative stress. Mitochondrion 2016; 30: 197–207.

54 Yuan N, Chen Y, Xia Y, Dai J, Liu C. Inflammation-related biomarkers in major psychiatric disorders: A cross-disorder assessment of reproducibility and specificity in 43 meta-analyses. Transl Psychiatry 2019; 9: 233.

55 Darrow SM, Verhoeven JE, Révész D, Lindqvist D, Penninx BWJH, Delucchi KL et al. The association between psychiatric disorders and telomere length: A meta-analysis involving 14,827 persons. Psychosom Med 2016; 78: 776–787.

56 Strawbridge R, Arnone D, Danese A, Papadopoulos A, Herane Vives A, Cleare AJ. Inflammation and clinical response to treatment in depression: A meta-analysis. Eur Neuropsychopharmacol 2015; 25: 1532–1543.

57 Moreno-Villanueva M, Kramer A, Hammes T, Venegas-Carro M, Thumm P, Bürkle A et al. Influence of acute exercise on DNA repair and PARP activity before and after irradiation in lymphocytes from trained and untrained individuals. Int J Mol Sci 2019; 20: 2999.

58 Kelly C, Trumpff C, Acosta C, Assuras S, Baker J, Basarrate S et al. A platform to map the mind–mitochondria connection and the hallmarks of psychobiology: the MiSBIE study. Trends Endocrinol Metab 2024; : S104327602400225X.

59 Junker A, Wang J, Gouspillou G, Ehinger JK, Elmér E, Sjövall F et al. Human studies of mitochondrial biology demonstrate an overall lack of binary sex differences: A multivariate meta-analysis. FASEB J 2022; 36. doi:10.1096/fj.202101628R.

60 Rausser S, Trumpff C, McGill MA, Junker A, Wang W, Ho S-H et al. Mitochondrial phenotypes in purified human immune cell subtypes and cell mixtures. eLife 2021; 10: e70899.

61 American Psychiatric Association. Diagnostic and Statistical Manual of Mental Disorders (DSM-5®). American Psychiatric Publishing: Washington, DC, 2013 10.1176/appi.books.9780890425596.

62 Schmahl C, Herpertz SC, Bertsch K, Ende G, Flor H, Kirsch P et al. Mechanisms of disturbed emotion processing and social interaction in borderline personality disorder: state of knowledge and research agenda of the German Clinical Research Unit. Borderline Personal Disord Emot Dysregulation 2014; 1: 12.

63 World Medical Association. Declaration of Helsinik: Ethical Principles for Medical Research Involving Human Subjects. J Am Med Assoc 2013; 310: 2191–2194.

64 Loranger AW, Sartorius N, Andreoli A, Berger P, Buchheim P, Channabasavanna SM et al. The International Personality Disorder Examination: The World Health Organization/Alcohol, Drug Abuse, and Mental Health Administration international pilot study of personality disorders. Arch Gen Psychiatry 1994; 51: 215–224.

65 Wittchen H-U, Zaudig M, Fydrich T. *Strukturiertes klinisches Interview für DSM-IV: SKID Achse I und II: Handanweisung*. Hogrefe: Göttingen, Germany, 1997.

66 Bohus M, Kleindienst N, Limberger MF, Stieglitz R-D, Domsalla M, Chapman AL et al. The Short Version of the Borderline Symptom List (BSL-23): Development and Initial Data on Psychometric Properties. Psychopathology 2009; 42: 32–39.

67 Zanarini MC. Zanarini rating scale for borderline personality disorder (ZAN-BPD): A continuous measure of DSM-IV borderline psychopathology. J Personal Disord 2003; 17: 233–242.

68 Klinitzke G, Romppel M, Häuser W, Brähler E, Glaesmer H. Die deutsche Version des Childhood Trauma Questionnaire (CTQ) – psychometrische Eigenschaften in einer bevölkerungsrepräsentativen Stichprobe. PPmP - Psychother · Psychosom · Med Psychol 2012; 62: 47–51.

69 Hautzinger M, Keller F, Kühner C. Beck Depressions-Inventar II (BDI II), Testhandbuch. Harcourt Test Services: Frankfurt a. M., Germany, 2006.

70 Laux L, Glanzmann P, Schaffner P, Spielberger CD. Das State-Trait-Angstinventar (STAI): theoretische Grundlagen und Handanweisung. Beltz: Weinheim, 1981 https://www.testzentrale.de/shop/das-state-trait-angstinventar.html (accessed 21 Jan2025).

71 Franke GH. SCL-90-R - Die Symptom-Checkliste von L. R. Derogatis. 2. vollständig überarbeitete und neu normierte Auflage. Beltz Test: Göttingen, 2002.

72 Reicherzer M, Brandl T. Der Fragebogen zu selbstverletzendem Verhalten (FSVV) – ein neues Erhebungsinstrument für die klinische Praxis. 2011.

73 Rohrmann S, Hodapp V, Schnell K, Tibubos AN, Schwenkmezger P, Spielberger CD. STAXI-2: das State-Trait-Ärgerausdrucks-Inventar-2; deutschsprachige Adaptation des State-Trait Anger Expression Inventory-2 (STAXI-2) von Charles D. Spielberger. Huber: Bern, 2013.

74 Werner R, Von Collani G. Deutscher Aggressionsfragebogen. Zusammenstellung Sozialwissenschaftlicher Items Skalen ZIS 2004. doi:10.6102/ZIS52.

75 Preuss UW, Rujescu D, Giegling I, Watzke S, Koller G, Zetzsche T et al. Psychometrische Evaluation der deutschsprachigen Version der Barratt-Impulsiveness-Skala. Nervenarzt 2008; 79: 305–319.

76 Ehring T, Svaldi J, Tuschen-Caffier B, Berking M. Validierung der Difficulties in Emotion Regulation Scale–deutsche Version (DERS-D). 2013.

77 Pesta D, Gnaiger E. High-resolution respirometry: OXPHOS protocols for human cells and permeabilized fibers from small biopsies of human muscle. Mitochondrial Bioenerg Methods Protoc 2012; : 25–58.

78 Larsen S, Nielsen J, Hansen CN, Nielsen LB, Wibrand F, Stride N et al. Biomarkers of mitochondrial content in skeletal muscle of healthy young human subjects. J Physiol 2012; 590: 3349–3360.

79 Eigentler A, Draxl A, Wiethüchter A, Kuznetsov A V., Lassing B, Gnaiger E. Laboratory protocol: citrate synthase, a mitochondrial marker enzyme. MiPNet 2012; 17: 1–11.

80 Singh NP, McCoy MT, Tice RR, Schneider EL. A simple technique for quantitation of low levels of DNA damage in individual cells. Exp Cell Res 1988; 175: 184–191.

81 Boeck C, Gumpp AM, Koenig AM, Radermacher P, Karabatsiakis A, Kolassa I-T. The association of childhood maltreatment with lipid peroxidation and DNA damage in postpartum women. Front Psychiatry 2019; 10. doi:10.3389/fpsyt.2019.00023.

82 Kumaravel TS, Vilhar B, Faux SP, Jha AN. Comet Assay measurements: a perspective. Cell Biol Toxicol 2009; 25: 53–64.

83 R Core Team. R: A language and environment for statistical computing. 2019.https://www.R-project.org/.

